# A clinical specific BERT developed with huge size of Japanese clinical narrative

**DOI:** 10.1101/2020.07.07.20148585

**Authors:** Yoshimasa Kawazoe, Daisaku Shibata, Emiko Shinohara, Eiji Aramaki, Kazuhiko Ohe

## Abstract

Generalized language models that pre-trained with a large corpus have achieved great performance on natural language tasks. While many pre-trained transformers for English are published, few models are available for Japanese text, especially in clinical medicine. In this work, we demonstrate a development of a clinical specific BERT model with a huge size of Japanese clinical narrative and evaluated it on the NTCIR-13 MedWeb that has pseudo-Twitter messages about medical concerns with eight labels. Approximately 120 millions of clinical text stored at the University of Tokyo Hospital were used as dataset. The BERT-base was pre-trained with the entire dataset and a vocabulary including 25,000 tokens. The pre-training was almost saturated at about 4 epochs, and the accuracies of Masked LM and Next Sentence Prediction were 0.773 and 0.975, respectively. The developed BERT tends to show higher performances on the MedWeb task than the other nonspecific BERTs, however, no significant differences were found. The advantage of training on domain-specific texts may become apparent in the more complex tasks on actual clinical text, and such corpus for the evaluation is required to be developed.

## Introduction

In recent years, generalized language models that perform pre-training on huge corpus have achieved great performance on a variety of natural language task. These language models are based on the transformer architecture that is a novel neural network based solely on a self-attention mechanism [1]. Some models such as Bidirectional Encoder Representations from Transformers (BERT) [2], Transformer-XL [3], XLNet [4], RoBERTa [5], XLM [6], GPT [7], GPT-2 [8] have been developed and updating state of the art results. It is considered that corpus for pre-training preferred to use the same domain as the target task. In the fields of life science or clinical medicine, domain specifically pre-trained model such as Sci-BERT [9], Bio-BERT [10], and Clinical-BERT [11], have been published for English texts. As for the Clinical-BERT, a study that domain specifically pre-training yields performance improvements on the tasks of common clinical Natural language processing (NLP) as compared to nonspecific model has been reported.

While many pre-trained transforms for English are published, few models are available for Japanese text, especially in clinical medicine. One of the options is to use multilingual BERT published by Google, however, the model would have disadvantage for word-based tasks because of its character-based vocabulary. As others for Japanese text, BERTs that have been pre-trained with Japanese WIKIPEDIA are published [12] [13], however, their applicability for the task of clinical medicine has not been studied. Because clinical narratives (physicians’ or nurses’ notes) should have differences in linguistic characteristics from text on the web, pre-training on clinical text would have an advantage for the clinical NLP tasks. In this work, we develop and publicly release a pre-trained BERT that make use of huge size of Japanese clinical narratives. We also demonstrate an evaluation with a shared NLP task to compare the developed clinical specific BERT with the three types of nonspecific BERTs for Japanese text.

## Methods

### Datasets

Approximately 120 million lines of clinical text for eight years stored at electronic health record system of the University of Tokyo Hospital were used. Data collection followed a protocol approved by the Institutional Review Board at the University of Tokyo Hospital (2019276NI). Those texts are mainly recorded by physicians and nurses for daily clinical practice. Because Japanese text includes two-byte full-width characters (mainly Kanji, Hiragana, or Katakana) and one-byte half-width characters (mainly ASCII characters), the Normalization Form Compatibility Composition (NFKC) followed by full-width characterization to all characters were applied as a pre-processing.

### Tokenization of Japanese text

To input a sentence into BERT, it is necessary to segment the sentence into tokens included in the BERT vocabulary. In non-segmented languages such as Japanese or Chinese, a tokenizer must accurately identify every word in a sentence before attempt to parse it and to do that requires a method of finding word boundaries without the aid of word delimiters. To obtain BERT tokens from a Japanese text, morphological analysis followed by wordpiece tokenization is applied. Morphological analyzer such as the MeCab [14] or the Juman++ [15] is commonly used in Japanese text processing to segment a source text into word units that are pre-defined in its own dictionary. Subsequently, wordpiece tokenization is applied, which segment a word unit into several pieces of tokens included in BERT vocabulary. In the wordpiece tokenization, the word like *playing* is segmented to *play* and *##ing*, the latter is called subword. Fig 1 shows a schematic view of morphological analysis and wordpiece tokenization of a Japanese text.

**Fig 1.**
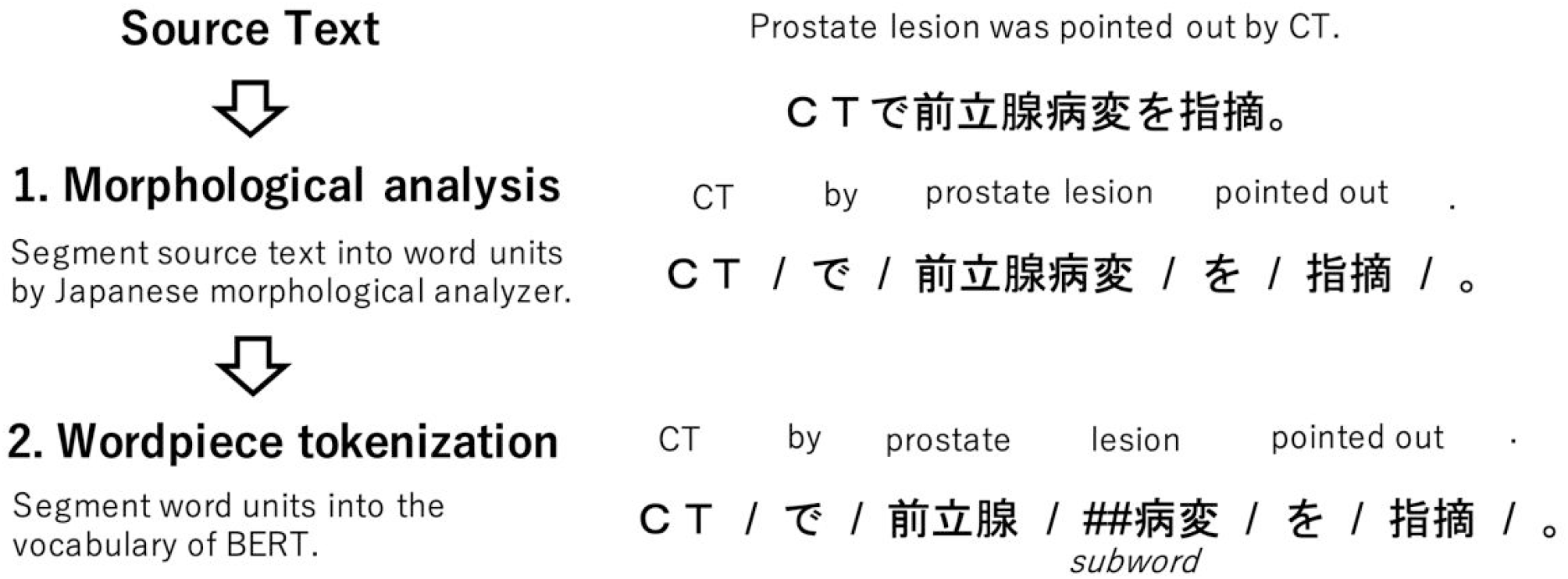
The schematic view of morphological analysis and wordpiece tokenization.

### Making BERT vocabulary

A BERT model requires a fixed number of token vocabulary for word pieces embeddings. To make the BERT vocabulary, candidate word pieces were obtained by applying morphological analysis followed by the Byte Pair Encoding (BPE) [16] to the entire dataset. The MeCab was used as a morphological analyzer along with the mecab-ipadic-NEologd [17] and the J-MeDic [18] as external dictionary. The former has been built utilizing various resources on the web, and it was used to identify personal names from the clinical text as much as possible and aggregate them into a special token (@@N). The latter is domain specific dictionary that has been built from Japanese clinical text, and it was used to segment words for disease or findings in as large a unit as possible. BPE first decompose a word unit into character symbols, and subsequently create a new symbol by merging two adjacent and highly frequent symbols. The merging process is stopped if the number of different symbols reaches the desired vocabulary size. In addition to this process, candidate words that evoke specific persons or facilities were excluded by manual screening, which make the developed BERT publicly available. Eventually, 25,000 tokens including special tokens were adopted as the vocabulary.

### Pre-training BERT

BERT has shown state of the art results for a wide range of tasks, such as single sentence classification, sentence pair classification, question answering without substantial task specific architecture modifications. The novelty of BERT is that it took an idea of learning word embeddings one step further, by learning each embedding vector directly from the sequence. In order to do this, BERT utilizes the self-attention mechanism, which learns sentence expressions by capturing co-occurrence relationships between words. Pre-training of BERT is performed by inputting fixed-length tokens obtained from two sentences and optimizing the Masked-LM and the Next Sentence Prediction simultaneously. Since these two tasks do not require manually supervised labels, the pre-learning is conducted as self-supervised learning.

#### Masked-LM

Fig 2A shows a schematic view of Masked-LM. This task masks, randomly replaces, or keeps each input token with a certain probability, and estimates the original embeddings of those tokens. By estimating not only the masked tokens but also the replaced or kept tokens, more appropriate representation of sentences is obtained. Although the selection probability of the tokens to be dealt with is arbitrary, we used the 15% mentioned in the original paper.

**Fig 2.**
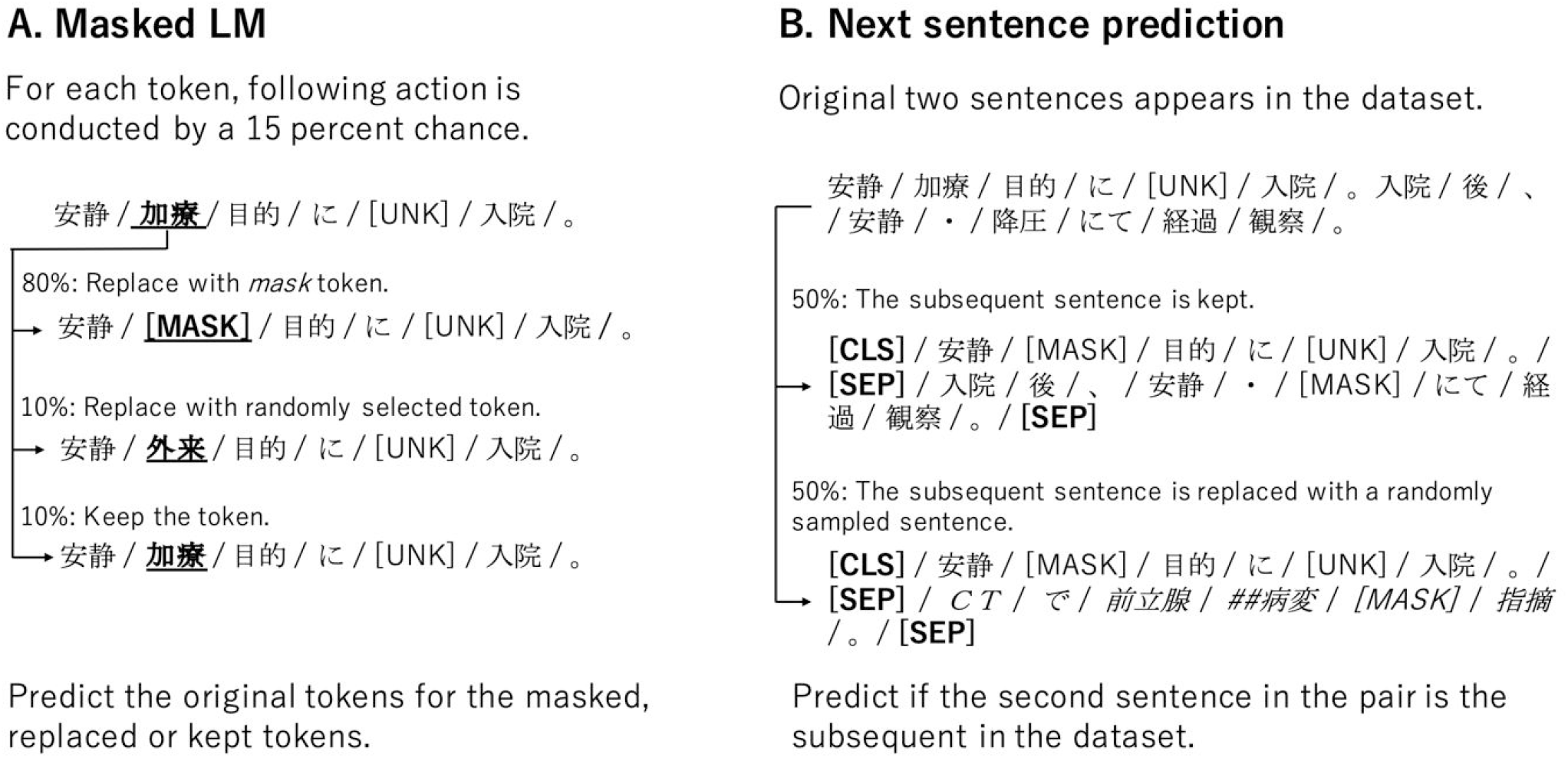
The schematic view of Masked-LM and Next Sentence Prediction task. **A**. Masked LM predicts the original tokens for the masked, replaced or kept tokens. **B**. Next Sentence Prediction predicts if the second sentence in the pair is the subsequent in the original documents. The role of special symbols are follows. [CLS] is added in front of every input text, and the output vector is used for Next Sentence Prediction task. [MASK] is masked token in Masked-LM task. [SEP] is a break between sentences. [UNK] is unknown token that does not appear in the vocabulary.

#### Next Sentence Prediction

Fig 2B shows a schematic view of Next Sentence Prediction. In this task, the model receives pairs of sentences and predicts whether the second sentence of the pair is the consecutive sentence in the original dataset. To develop such a training dataset, for two consecutive sentences in the original dataset, connect the original consecutive sentences with a probability of 50% as a positive example, and connect the remaining 50% of the sentences randomly sampled as a negative example. In general, two sentences connected by a period are assumed to be consecutive, and two sentences that pinch a line feed code are assumed to be non-consecutive. However, because for simply displaying clinical narratives in a narrow screen area, it seemed to be that clinical sentences tend to pinch a line feed code even if they were originally consecutive sentences. In other words, a series of sentences referring to the same topic appears as a list of non-consecutive short sentences, which is considered to have a negative impact on the training of Next Sentence Prediction. For this reason, we treated all sentences appearing in a document recorded in one day for a patient as consecutive sentences.

### Evaluation task

The performance of the developed BERT was evaluated by fine-tuned approach using the NTCIR-13 Medical Natural Language Processing for Web Document (MedWeb) [19]. The MedWeb is publicly available and provides pseudo-Twitter messages about medical concerns in a cross-language and multi-label corpus, covering three languages (Japanese, English, and Chinese), and annotated with eight labels. Positive or Negative is given to eight labels of Influenza, Diarrhea, Hay fever, Cough, Headache, Fever, Runny nose, and Cold, and Positive may be given to multiple labels in a message. We performed a multi-label task to classify these eight classes simultaneously. Table 1 shows examples of each set of pseudo-tweets.

**Table 1.**
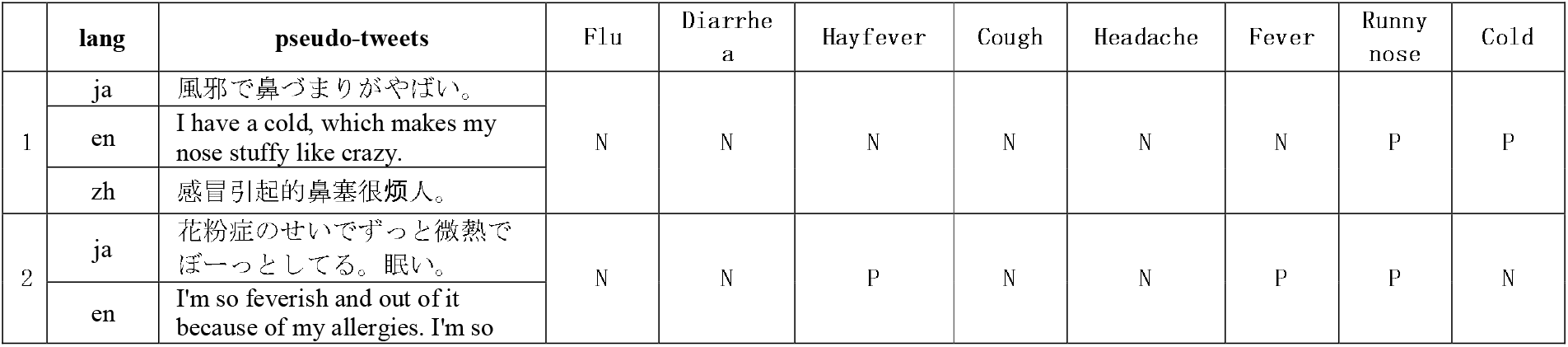
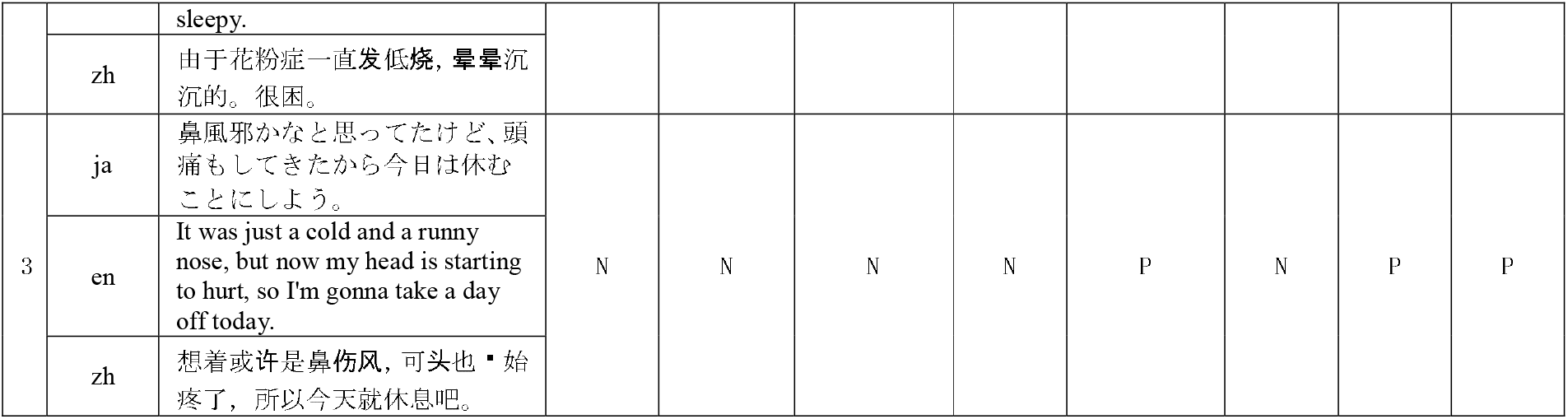
Three examples of pseudo-tweets with the eight class of symptoms. English sentence (en) and Chinese sentence (zh) were translated from Japanese sentence (ja).

### Experiment settings

For the pre-training experiments, we leverage the Tensorflow implementation of BERT-BASE (12 layers, 12 attention heads, 768 embedding dimension, 110 million parameters) published by Google [2]. Approximately 99% of 120 million sentences was used for training, and the remaining of 1% was used for the evaluation for the accuracies of Masked LM and Next Sentence Prediction.

For the evaluation experiments, the pre-trained BERT was fine-tuned. The network was configured such that the output vector C corresponding to the first input token ([CLS]) is linearly transformed to eight labels by a fully connected layer, and the positive or negative of each of the eight labels are outputted through sigmoid function. Binary cross entropy was used for the loss function, and the learning rate was optimized by Adam initialized with 1e-5. All network parameters including BERT are updated during this fine-tuning process. Fig 3 shows a schematic view of this network. The models are trained and tuned by five-folds cross-validation, which split 1,920 MedWeb training data into 4:1. The average results of each five tuned model for 640 MedWeb test data was assessed. The performance was assessed by the exact match accuracy and label-wise F-measure (macro F1).

**Fig 3.**
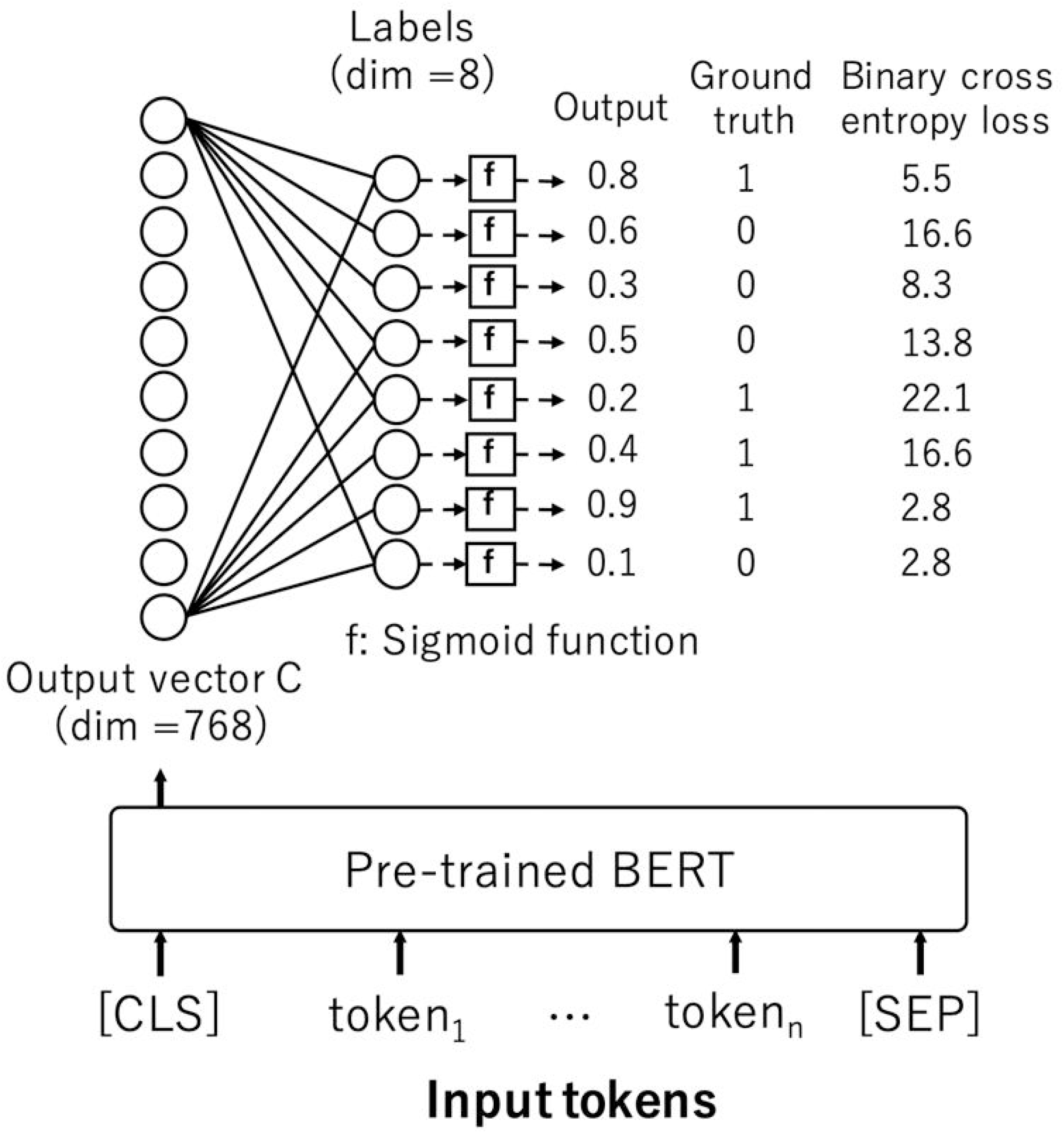
The schematic view of the network for evaluation.

To inspect an advantage of the domain specific model, we also evaluated the two kinds of domain nonspecific BERT that are pre-trained in Japanese Wikipedia and Google multilingual BERT. In addition, to inspect the difference of the performance due to input features, we also evaluated Support Vector Machine (SVM) and Logistic Regression (LR) that use the same morphological analyzer, dictionary and vocabulary to each BERT. Table 2 shows the specifications of each BERT model. In this table, the total number of unknown tokens appeared in the MedWeb dataset is shown, which are obtained by the tokenizer with each BERT vocabulary.

**Table 2.**
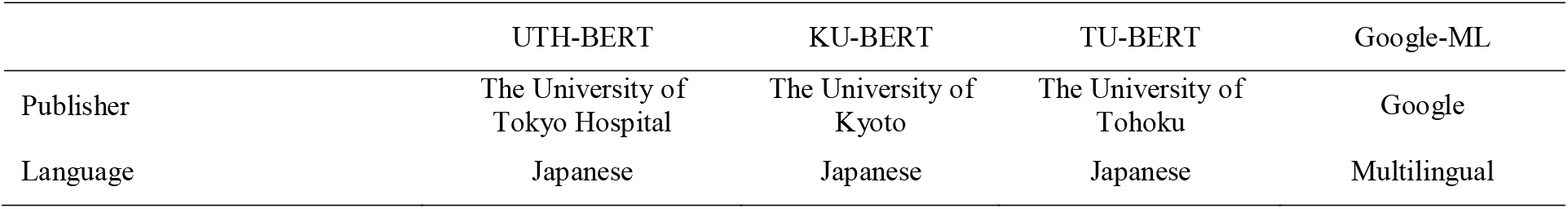
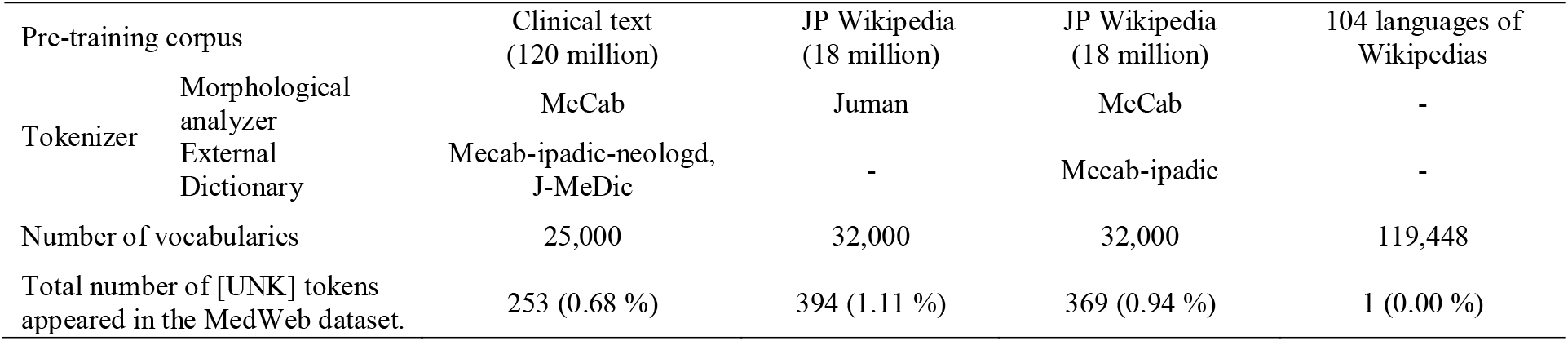
The specifications of each BERT

## Results

### Pre-training performance

Table 3 shows the results of the pre-training. The pre-training was almost saturated at about 10 million steps (4 epochs), and the accuracies of Masked LM and Next Sentence Prediction were 0.773 and 0.975, respectively. With a mini-batch size of 50, 2.5 million steps are equivalent to approximately 1 epoch. It took about 45 days to learn 4 epochs using single GPU. In the following experiment, UTH-BERT with 10 million steps of training was used.

**Table 3.**
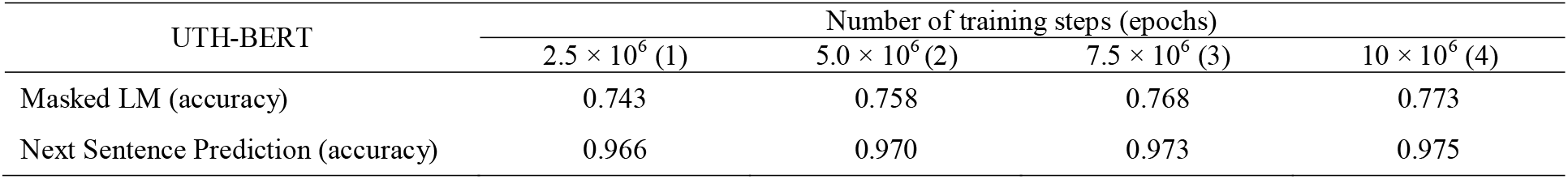
Accuracies of Masked LM and Next Sentence Prediction in pre-training for the evaluation dataset.

### Finetuning performance

Table 4 shows exact match performance of four kind of pre-trained BERT, SVM and LR. Although there was no significant difference due to the number of cross validations, the average performances have better tendency in the order of BERT models (0.851), LR models (0.724) and SVM models (0.701). Among the BERT models, UTH-BERT (0.862) performed the best and Google-ML BERT (0.827) performed the worst. There were slight differences among UTH-BERT (0.862), KU-BERT (0.855) and TU-BERT (0.858), which indicates no significant evidence for the advantage of domain-specific model. For SVM models, the performances have better tendency in the order of SVM1, SVM2, SVM3, which use the same tokenizer and vocabulary to UTH-BERT, KU-BERT and TU-BERT, respectively. This considered to indicate the suitability of each vocabulary for this evaluation task. It was also that LR models showed the same tendency as SVM models.

**Table 4.**
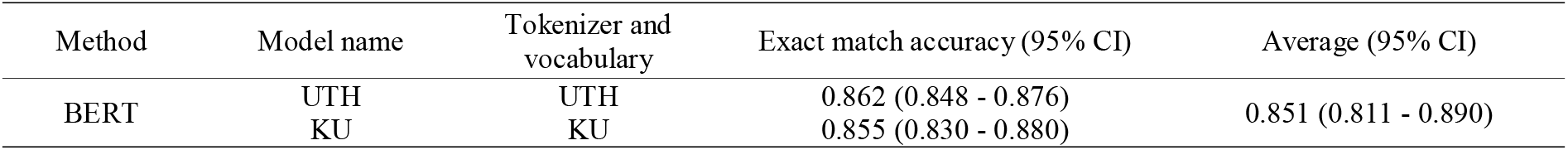
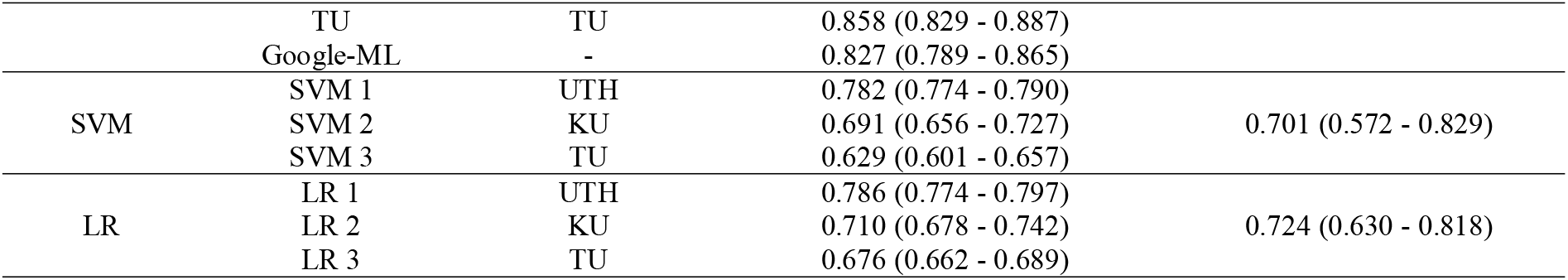
The exact match accuracy of each model by the five-folds cross validation.

Table 5 shows the label-wise F-measure (macro F1) of each model. Like the exact match performance, the average F-measures have better tendency in the order of BERTs (0.883), LRs (0.839), and SVMs (0.827), and the average performances of the eight classes among BERTs were highest for UTH-BERT (0.893) and lowest for Google-ML BERT (0.867). As for the performances for each class, the performances of SVM2 (0.576), SVM3 (0.507), and LR2 (0.610), LR3 (0.539) for *cough* were significantly lower than the others (95% CIs were not shown).

**Table 5.** The label-wise performances of each model by the five-folds cross validation. The results are shown as F-measure (macro F1).

| Method | Model | Tokenizer and vocabulary | class |  |  |  |  |  |  |  | Ave.in class (95% CI) | Ave. in methods (95% CI) |
| --- | --- | --- | --- | --- | --- | --- | --- | --- | --- | --- | --- | --- |
|  |  |  | *flu* | *Diarrhea* | *Hay fever* | *Cough* | *Headache* | *Fever* | *Runny nose* | *Cold* |  |  |
| BERT | UTH | UTH | 0.747 | 0.929 | 0.880 | 0.940 | 0.960 | 0.851 | 0.933 | 0.901 | 0.893 (0.880 - 0.905) | 0.883 (0.865 - 0.900) |
|  | KU | KU | 0.708 | 0.923 | 0.873 | 0.945 | 0.946 | 0.841 | 0.921 | 0.924 | 0.885 (0.869 - 0.901) |  |
|  | TU | TU | 0.706 | 0.934 | 0.870 | 0.948 | 0.951 | 0.843 | 0.929 | 0.900 | 0.885 (0.867 - 0.903) |  |
|  | Google-ML | - | 0.659 | 0.893 | 0.872 | 0.937 | 0.956 | 0.806 | 0.912 | 0.904 | 0.867 (0.850 - 0.885) |  |
| SVM | SVM 1 | UTH | 0.717 | 0.888 | 0.871 | 0.932 | 0.883 | 0.787 | 0.881 | 0.891 | 0.856 (0.842 - 0.870) | 0.827 (0.794 - 0.860) |
|  | SVM 2 | KU | 0.703 | 0.909 | 0.859 | 0.576 | 0.964 | 0.810 | 0.881 | 0.878 | 0.823 (0.811 - 0.834) |  |
|  | SVM 3 | TU | 0.689 | 0.901 | 0.847 | 0.507 | 0.949 | 0.778 | 0.890 | 0.867 | 0.804 (0.770 - 0.836) |  |
| LR | LR 1 | UTH | 0.764 | 0.866 | 0.882 | 0.923 | 0.872 | 0.766 | 0.895 | 0.899 | 0.858 (0.839 - 0.878) | 0.839 (0.810 - 0.869) |
|  | LR 2 | KU | 0.747 | 0.894 | 0.893 | 0.610 | 0.946 | 0.792 | 0.903 | 0.902 | 0.836 (0.813 - 0.859) |  |
|  | LR 3 | TU | 0.729 | 0.904 | 0.873 | 0.539 | 0.943 | 0.800 | 0.918 | 0.881 | 0.823 (0.794 - 0.853) |  |
| Ave. |  |  | 0.717 | 0.904 | 0.872 | 0.786 | 0.937 | 0.807 | 0.906 | 0.895 |  |  |

## Discussions

### Interpretations of BERT model

In this study, we demonstrate a pre-training of BERT model with a huge size of Japanese clinical narrative and evaluated it on the NTCIR-13 MedWeb task. To the best of our knowledge, this work is the first to publish and inspect a BERT model that is pre-trained by Japanese text in clinical domain. Among the machine learning methods, BERT shows the most successful performance both in the exact match accuracy and the label-wise F-measure. A possible explanation for the success is that it is pre-trained on a large corpus, which affords good representation of the input sentence. As we shown in Table 5, the performances of SVM2, SVM3 and LR2, LR3 for *cough* were significantly lower than the others. This is because the vocabulary of KU-BERT for SVM2 and LR2, and TU-BERT for SVM3 and LR3 have few variations of *cough* symbols (zero in KU-BERT, two in TU-BERT and fifteen in UTH-BERT) and the relevant words tend to be treated as unknown words. Those unknown words are semantically ambiguous and had a negative impact on the classifier. Nevertheless, the performances of KU-BERT and TU-BERT for *cough* did not decrease. This was considered to be due to the Masked-LM that acquires sentence expressions from the context even if there are some unknown words. Similar phenomenon was also observed in SVM and LR models in Table 4. In this table, SVM 2, SVM 3, LR2 and LR3 have significantly lower performance than SVM 1 or LR1, which indicate less adaptable of the vocabulary of KU-BERT and TU-BERT for this evaluation task. However, KU-BERT and TU-BERT themselves show comparable performance to UTHE-BERT. This indicate that even if the vocabulary is less adaptable to the evaluation task, the negative impact on BERT performance is not so strong.

### Advantage of pre-training on domain text

Among the BERT models, the performances of UTH-BERT, KU-BERT, and TU-BERT, which specialized to Japanese texts showed higher tendency than Google multilingual BERT. The multilingual BERT use character-based vocabulary that alleviate vocabulary problems for handling multiple languages instead of giving up the semantic information that words have. This result would indicate a disadvantage of character-based vocabulary compared to word-based vocabulary. Even so, the performances of the multilingual BERT were comparable to the other Japanese specific BERT models. With regard to the advantage of pre-training with domain-specific text, UTH-BERT pre-trained with clinical text tended to show higher performances than KU-BERT and TU-BERT pre-trained with Wikipedia, however, no significant differences were found. One of the reasons is that because sentence classification is a relatively easy task for BERTs which have pre-trained on large text, the advantage of pre-learning with the domain text may not have been noticeable. Another reason is that because the NTCIR-13 MedWeb used for the evaluation is intermediate corpus between WEB and medicine, the differences among BERTs may not have been clear. The advantage of training on domain-specific texts may become apparent in the more complex tasks such as question answering or causality recognition using actual clinical text, and such corpus for the evaluation is required to be developed.

### Limitations and future works

Given that our developed BERT was evaluated exclusively on the NTCIR-13 MedWeb task, there is currently a limitation in the generalizability of the performance. Our purpose is to develop tools that will be useful for NLP in clinical domain, though, knowing their nature requires evaluations on the more complex clinical or non-clinical tasks. In parallel, error analysis is required to investigate the mistakes made by the BERT based model. For this purpose, visualizing self-attentions [20] in the BERT model will be useful. It is also that useful linguistic information can be found not only in the self-attentions themselves, but also in the attention maps that represent the relation of word to word attention between adjacent layers [21]. In future task, the detailed error analysis with those visualizing method of self-attentions mechanism is required, which may contribute in the understanding of how BERT models understand language.

## Data Availability

1. Clinical text used for pre-training the UTH-BERT cannot be shared because of not publicly available, and restrictions apply to their use.
2. The UTH-BERT model is available for non-commercial use at the following URL: https://ai-health.m.u-tokyo.ac.jp/uth-bert
3. NTCIR-13 dataset is available upon request at the following URL: https://www.nii.ac.jp/dsc/idr/ntcir/ntcir.html

https://www.nii.ac.jp/dsc/idr/ntcir/ntcir.html

## Supporting information

The pre-trained BERT model with instruction for running the code is available below. https://ai-health.m.u-tokyo.ac.jp/uth-bert

## Author contributions

Conceptualization: YK, ES

Investigation: DS

Methodology: YK, DS

Resources: ES, KO

Supervision: EA, KO

Writing - original draft: YK

Writing - review & editing: YK, EA

## References

1. Ashish Vaswani, Noam Shazeer, Niki Parmar, Jakob Uszkoreit, Llion Jones, Aidan N Gomez, Łukasz Kaiser, Illia Polosukhin. Attention is all you need. Advances in neural information processing systems, pp.5998–6008, 2017.

2. Jacob Devlin, Ming-Wei Chang, Kenton Lee, Kristina Toutanova. BERT: Pre-training of Deep Bidirectional Transformers for Language Understanding. arXiv, 2018; ArXiv: 1810.04805.

3. Zihang Dai, Zhilin Yang, Yiming Yang, Jaime Carbonell, Quoc V. Le, Ruslan Salakhutdinov. Transformer-XL: Attentive Language Models Beyond a Fixed-Length Context. arXiv, 2019; ArXiv: 1901.02860.

4. Zhilin Yang, Zihang Dai, Yiming Yang, Jaime Carbonell, Ruslan Salakhutdinov, Quoc V. Le. XLNet: Generalized Autoregressive pretraining for Language Understanding. arXiv, 2019; ArXiv: 1906.08237.

5. Yinhan Liu, Myle Ott, Naman Goyal, Jingfei Du, Mandar Joshi, Danqi Chen, Omer Levy, Mike Lewis, Luke Zettlemoyer, Veselin Stoyanov. RoBERTa: A Robustly Optimized BERT pretraining Approach. arXiv, 2019; 1907.11692.

6. Guillaume Lample, Alexis Conneau. Cross-lingual Language Model pretraining. Advances in neural information processing systems, pp.7059–7069, 2019.

7. Alec Radford, Karthik Narasimhan, Tim Salimans, and Ilya Sutskever. Improving language understanding by generative Pre-training. OpenAI Blog, 2018.

8. Alec Radford, Jeffrey Wu, Rewon Child, David Luan, Dario Amodei, and Ilya Sutskever. Language models are unsupervised multitask learners. OpenAI Blog, 2019.

9. Iz Beltagy, Kyle Lo, Arman Cohan. SciBERT: A Pre-trained Language Model for Scientific Text. arXiv, 2019; ArXiv: 1903.10676.

10. Jinhyuk Lee, Wonjin Yoon, Sungdong Kim, Donghyeon Kim, Sunkyu Kim, Chan Ho So, Jaewoo Kang. BioBERT: a pre-trained biomedical language representation model for biomedical text mining. arXiv, 2019; ArXiv: 1901.08746.

11. Emily Alsentzer, John R. Murphy, Willie Boag, Wei-Hung Weng, Di Jin, Tristan Naumann, Matthew B. A. McDermott. Publicly Available Clinical BERT Embeddings. arXiv, 2019; ArXiv: 1904.03323.

12. a BERT published by the Kyoto University. http://nlp.ist.i.kyoto-u.ac.jp/EN/.(cited 2020-March-9).

13. a BERT published by the Tohoku University. https://github.com/cl-tohoku/bert-japanese. (xcited 2020-March-9).

14. Kudou Taku. MeCab : Yet Another Part-of-Speech and Morphological Analyzer (in Japanese). https://github.com/taku910/mecab.

15. Daisuke Kawahara, Sadao Kurohashi. A Fully-Lexicalized Probabilistic Model for Japanese Syntactic and Case Structure Analysis, In Proceedings of the Human Language Technology Conference of the North American Chapter of the Association for Computational Linguistics (HLT-NAACL2006), pp.176–183, 2006.

16. Rico Sennrich, Barry Haddow, Alexandra Birch. Neural Machine Translation of Rare Words with Subword Units. Proceedings of the 54th Annual Meeting of the Association for Computational Linguistics. pp.1715–1725, 2016.

17. Toshinori Sato, Taiichi Hashimoto, Manabu Okumura. Implementation of a word segmentation dictionary called mecab-ipadic-NEologd and study on how to use it effectively for information retrieval (in Japanese). Proceedings of the Twenty-three Annual Meeting of the Association for Natural Language Processing, NLP2017-B6-1, 2017.

18. Kaoru Ito, Hiroyuki Nagai, Taro Okahisa, Shoko Wakamiya, Tomohide Iwao, Eiji Aramaki. J-MeDic: A Japanese Disease Name Dictionary based on Real Clinical Usage. Proceedings of the Eleventh International Conference on Language Resources and Evaluation, 2018.

19. Shoko Wakamiya, Mizuki Morita, Yoshinobu Kano, Tomoko Ohkuma, Eiji Aramaki: Overview of the NTCIR-13 MedWeb Task, Proceedings of the 13th NTCIR Conference on Evaluation of Information Access Technologies, pp.40–49, 2017.

20. Jesse Vig. Visualizing Attention in Transformer-Based Language Representation Models. arXiv, 2019; ArXiv: 1904.02679.

21. Kevin Clark, Urvashi Khandelwal, Omer Levy, Christopher D. Manning. What Does BERT Look At? An Analysis of BERT’s Attention. arXiv, 2019; 1906.04341.

